# Comparison of infectious SARS-CoV-2 from the nasopharynx of vaccinated and unvaccinated individuals

**DOI:** 10.1101/2021.12.28.21268460

**Authors:** Mario A. Peña-Hernández, Jon Klein, Amyn A. Malik, Andreas Coppi, Chaney Kalinich, Chantal B. F. Vogels, Julio Silva, Yale SARS-CoV-2 Genomic Surveillance Initiative, David R. Peaper, Marie-Louise Landry, Craig Wilen, Nathan D. Grubaugh, Wade Schulz, Saad B. Omer, Akiko Iwasaki

## Abstract

The frequency of SARS-CoV-2 breakthrough infections in fully vaccinated individuals increased with the emergence of the Delta variant, particularly with longer time from vaccine completion. However, whether breakthrough infections lead to onward transmission remains unclear. Here, we conducted a study involving 125 patients comprised of 72 vaccinated and 53 unvaccinated individuals, to assess the levels of infectious virus in vaccinated and unvaccinated individuals. Quantitative plaque assays showed no significant differences in the titers of virus between these cohorts. However, the proportion of nasopharyngeal samples with culturable virus was lower in the vaccinated patients relative to unvaccinated patients (21% vs. 40%). Finally, time-to-event analysis with Kaplan-Myer curves revealed that protection from culturable infectious virus waned significantly starting at 5 months after completing a 2-dose regimen of mRNA vaccines. These results have important implications in timing of booster dose to prevent onward transmission from breakthrough cases.

---

Vaccines encoding Spike protein are highly effective in limiting the development of severe COVID-19 [1]; however, the durability of this protection against infection with heterologous SARS-CoV-2 Spike variants following standard vaccine regimens is currently unclear. The frequency of ‘breakthrough’ infections in fully vaccinated individuals increased with the appearance of the Delta variant (lineages B.1.617.2 and AY.3), suggesting that novel viral variants may mediate the degree of escape from immunity within vaccinated individuals [2, 3]. Previous studies have also demonstrated that breakthrough infections with Delta generate similar levels of viral nucleic acids among vaccinated and unvaccinated individuals [4, 5]. However, it is unclear whether there are differences in infectivity between vaccinated and unvaccinated individuals with similar Ct values. In addition, the durability of protection afforded by a 2-dose vaccination regimen against the presence of infectious virus in vaccinated individuals infected with SARS-CoV-2 variants remains unknown.

To address this question, we conducted a study involving 125 patients comprised of 72 vaccinated and 53 unvaccinated individuals. Nasopharyngeal samples from infected patients were obtained from excess clinical samples used for routine diagnosis of SARS-CoV-2 infection within the Yale New Haven Health (YNHH) system. Our study collected samples from July to August 2021, during which Delta was the predominant circulating SARS-CoV-2 variant within our region [6]. Nasopharyngeal samples from were selected from a range of RT-qPCR cycle threshold (Ct) values among vaccinated individuals and then matched with corresponding Ct values among unvaccinated individuals (Ct 13-20: 39% | 43%; Ct 21-30: 35% | 41%; Ct >30: 26% | 16%). Within the previously mentioned Ct ranges, patients were matched by age and sex to control for any demographic differences between cohorts. After generating matched vaccinated and unvaccinated cohorts, we confirmed there were no significant differences in age or sex (Mean age: 46.9 years, IQR: 29.8 – 60.6 years; 41.6% male). We also confirmed there were no differences in their time after symptom onset (range from 0-11 days and an average of 1.8 and 1.7 days for unvaccinated and vaccinated cohorts, respectively). Among the vaccinated cohort, 65 of 72 individuals who had breakthrough infections completed a standard two-dose mRNA vaccination program - either BNT162b2 or mRNA-1273 - with an average time between completion of mRNA vaccination series and breakthrough infection of 121.8 days (IQR: 83.7 – 161.0 days). 7 of 72 vaccinated individuals in our cohort received a single dose of a SARS-CoV-2 adenovirus vectored vaccine (Johnson and Johnson).

We observed no significant difference in SARS-CoV-2 genome copy numbers between vaccinated and unvaccinated infections after correcting for differences in clinical testing platforms (Figure 1A, Extended Methods). To assess whether there were differences in levels of infectious virus between breakthrough infections and unvaccinated individuals, we performed quantitative plaque assays and found no significant differences between cohorts (Figure 1B). We next analyzed the proportion of nasopharyngeal samples with culturable virus and found a lower percentage of infectious samples in vaccinated patients relative to unvaccinated patients (21% vs. 40%, RR: .53 (.30 - .91); Koopman asymptomatic score) (Figure 1C). Collectively, these observations suggest that SARS-CoV-2 vaccines may not alter the levels of replicating virus in the upper respiratory tract once viral infection establishes but may instead protect through accelerated clearance or enhanced neutralization of infectious viral particles [6].

**Figure 1.**
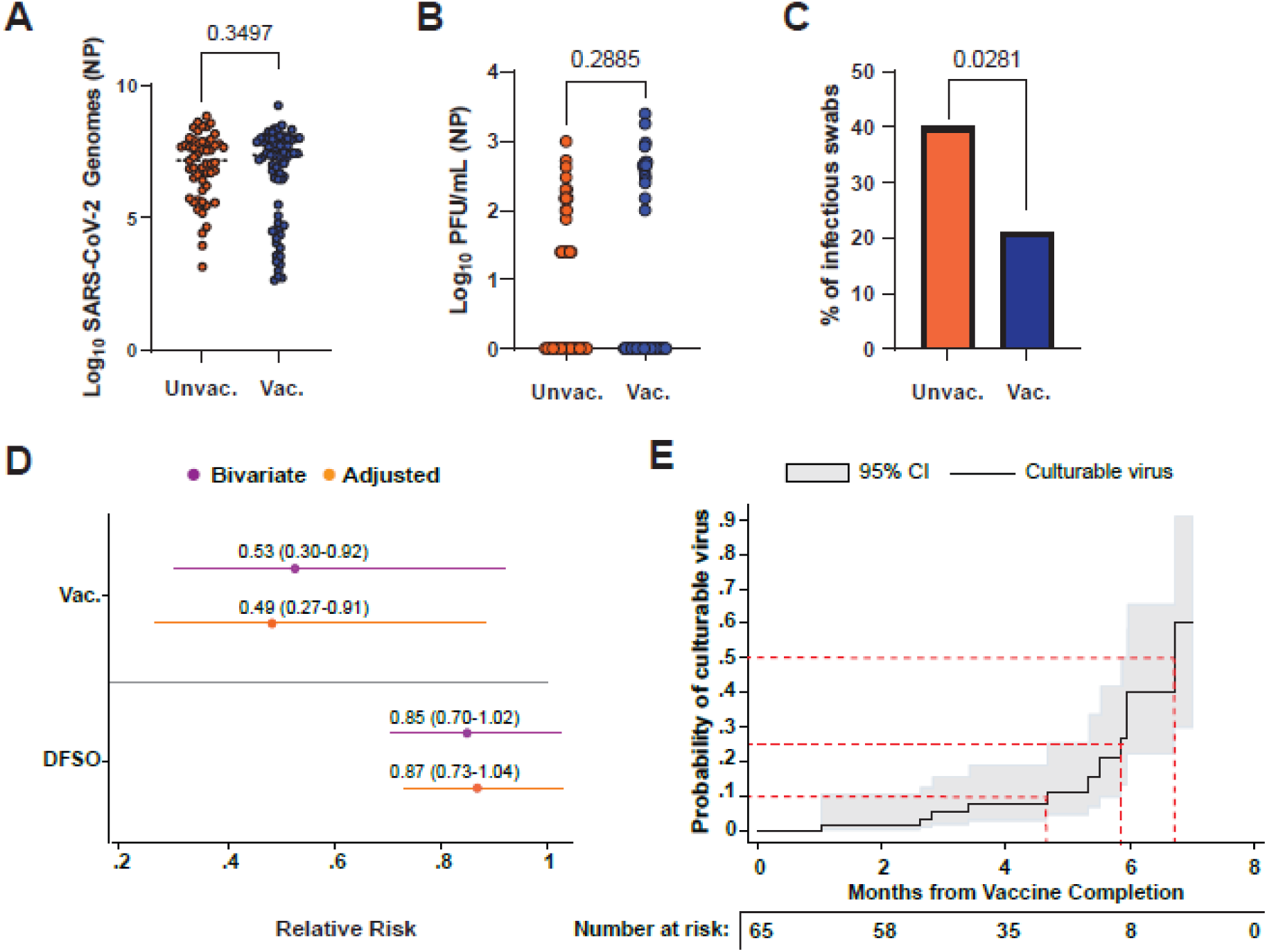
Isolation of infectious SARS-CoV-2 from vaccinated and unvaccinated individuals reveals waning protection beginning at 5 months. (**A**) Violin plots of log10 transformed SARS-CoV-2 viral genomes from nasopharyngeal swabs from vaccinated and unvaccinated individuals. Copy number estimates of SARS-CoV-2 viral genomes were performed using linear regressions to correct for differences in clinical testing platforms and nucleic acid isolation procedures (Extended Methods). Solid lines represent the median values, and dashed lines represent quartiles within each group. Significance was assessed using a two-tailed, unpaired Student’s t-test with Welch’s correction for unequal variances (t=0.9388, df=122.8). Exact p-values are reported. (**B**) Violin plots of log10 transformed particle forming units / mL (PFU / mL) of infectious virus isolated from nasopharyngeal swabs from vaccinated and unvaccinated individuals. Solid lines represent the median values, and dashed lines represent quartiles within each group. Significance was assessed using a two-tailed, unpaired Student’s t-test (t=1.066, df=123). Exact p-values are reported. (**C**) Proportion of swabs with infectious virus for unvaccinated and vaccinated individuals. Significance was assessed using Fisher’s Exact test and P-value threshold is reported. (**D**) Relative risk of isolating culturable virus amongst vaccinated and unvaccinated individuals with COVID-19 infection and the 95% confidence intervals in parenthesis. Significance was assessed using robust standard errors (**E**) Probability of culturable virus from time to completion of vaccination (black) along with respective 95% confidence intervals (grey) amongst those with completed mRNA vaccination schedules. Red dashed lines indicate fixed levels of probability of culturing infectious virus (10%, 25%, and 50%) with corresponding months since vaccine completion. *Abbreviations (left to right): NP = nasopharyngeal; Unvac. = Unvaccinated; Vac. = Vaccinated; PFU = plaque forming units; DFSO = Days from symptom onset; CI = Confidence Interval*.

To address whether the probability of culturing infectious virus depended on individual demographic factors or differences in COVID-19 disease trajectories, we used a log-binomial model with robust standard errors to predict probability of infectious virus among vaccinated and unvaccinated individuals. After accounting for differences in age, sex, and relative days from symptom onset, we found that vaccination significantly reduced the presence of infectious virus during breakthrough infection by 51% (aRR: 0.49, 95%CI: 0.27-0.91; Figure 1D). Among patients that completed 2 doses of the mRNA vaccine (65 of 72 vaccinated patients), time-to-event analysis with Kaplan-Myer curves revealed this protection waned significantly with time since vaccination. Investigations into various windows of time since completion of vaccination schedules indicated a progressive loss of protection beginning at 5 months, where the probability of isolating infectious virus was estimated at 11.0% (95% CI: 4.5-25.4). This rapidly increased to approximately 40.3% (95% CI: 22.0-65.6) by 6 months following vaccine completion (Figure 1E), suggesting substantial losses in protection against the presence of infectious virus among vaccinated individuals.

These results agree with previous studies that suggest mRNA vaccination protection decreases over time [7, 8], and also further suggests the broad adoption of mRNA boosters in a critical 4–6-month window may reduce transmissibility of SARS-CoV-2 among fully vaccinated individuals with breakthrough infections.

## METHODS

### Sample collection and quantification of infectious viral load

Nasopharyngeal swabs were collected by trained clinical personnel and swabs stored and frozen in universal viral transport media (VTM) prior to plaquing. SARS-CoV-2 positive vaccinated individuals were selected from the Yale New Heaven Health sytem within the period of 07/06/21 - 08/13/21. qRT-PCR cycle thresholds values ranged from 13-40. Unvaccinated individuals’ samples were age, sex, and Ct matched to the vaccinated samples.

SARS-CoV-2 titers from nasopharyngeal swabs were assessed by conventional plaque assay. Each nasopharyngeal sample was ten-fold serially diluted (dilution range from 1:10 to 1:1000000), added on a monolayer of Vero E6 cells, and fixed and stained at 72 hours post inoculation with 4% Paraformaldehyde and 0.1% of crystal violet respectively.

### PCR cycle threshold (Ct) correction and virus genome copy calculation

Diagnostic PCR Ct values for SARS-CoV-2 nasal swab samples were from 8 different testing platforms. In order to compare viral loads between different laboratories, testing method, and/or machines, we performed linear regression on a separate data set of SARS-CoV-2 RT-qPCR data from specimens tested the same platforms and subsequently tested for research and public health purposes by the Yale SARS-CoV-2 Genomic Surveillance Initiative. 7 of 8 platforms, including 119/125 specimens in this data set, had adequate data to correct. In order to transform these Cts normalized across platforms into genome equivalents, RNA transcripts corresponding to a segment of the SARS-CoV-2 nucleocapsid gene were serially diluted from 10^6^ to 10^0^ and evaluated in duplicate on the research RT-qPCR test. The best-fit linear regression of the average Ct on the log10-transformed standard values had slope 03.60971 and intercept 40.93733 (R^2^ = 0.99). Code can be found at https://github.com/cck42/delta-breakthrough.

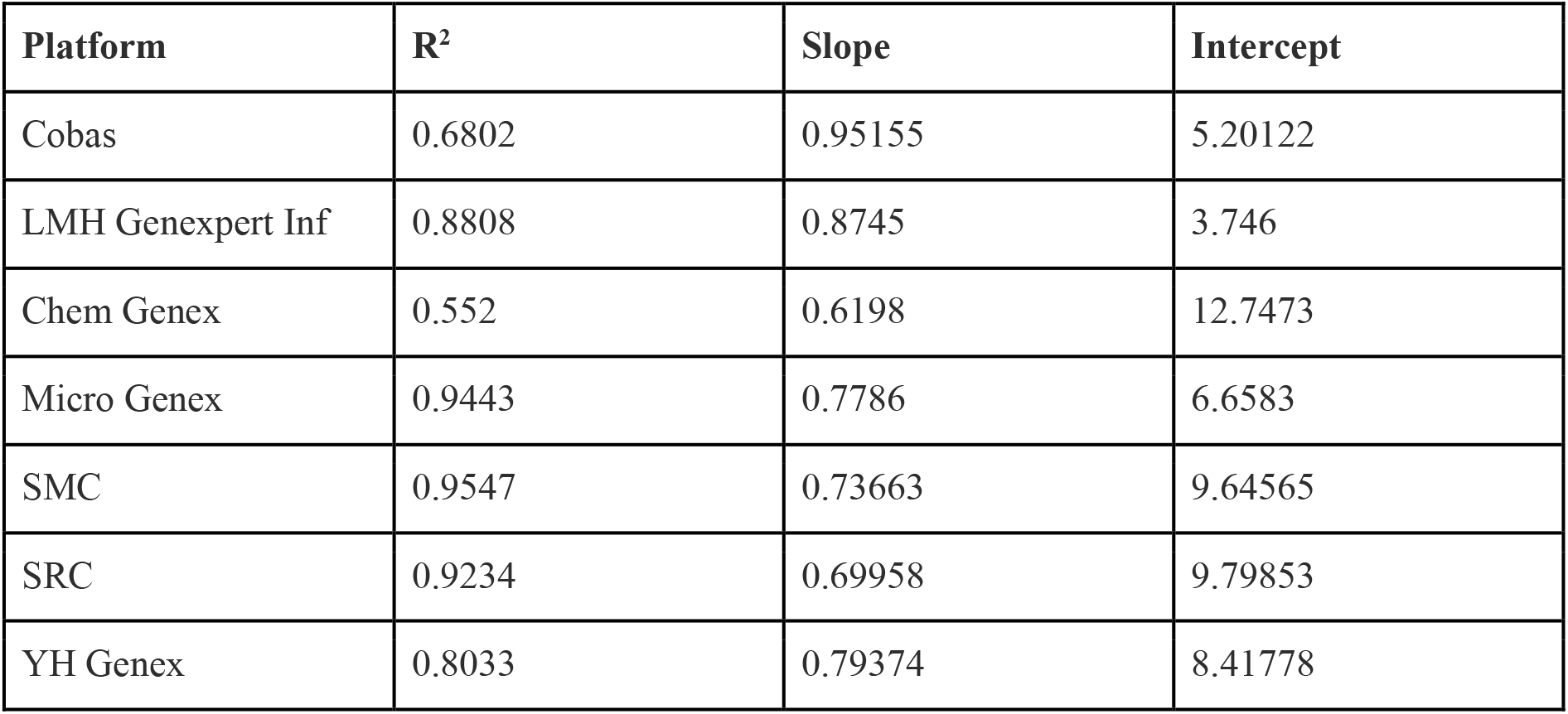

## Data Availability

All data produced in the present work are contained in the manuscript.

## Supplementary Figures

**Supplemental figure 1.**
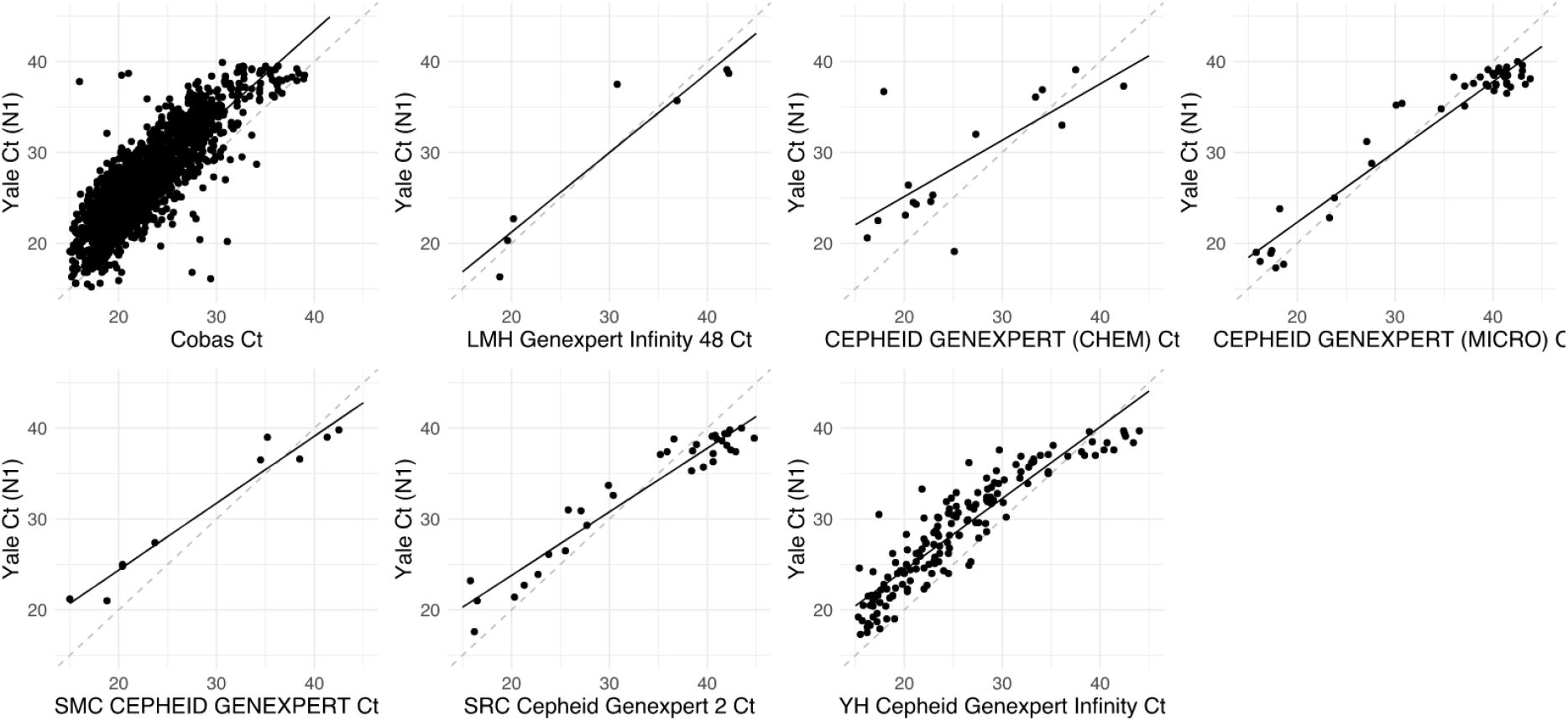
Ct values across platforms. Points represent Ct values of specimens tested both on Yale-New Haven Hospital diagnostic platforms and subsequently by the Yale SARS-CoV-2 Genomic Surveillance Initiative. Solid black line represents the line of best fit, and dashed gray line represents 1:1 line that would be expected if labs produced identical results.[9]

**Supplemental figure 2.**
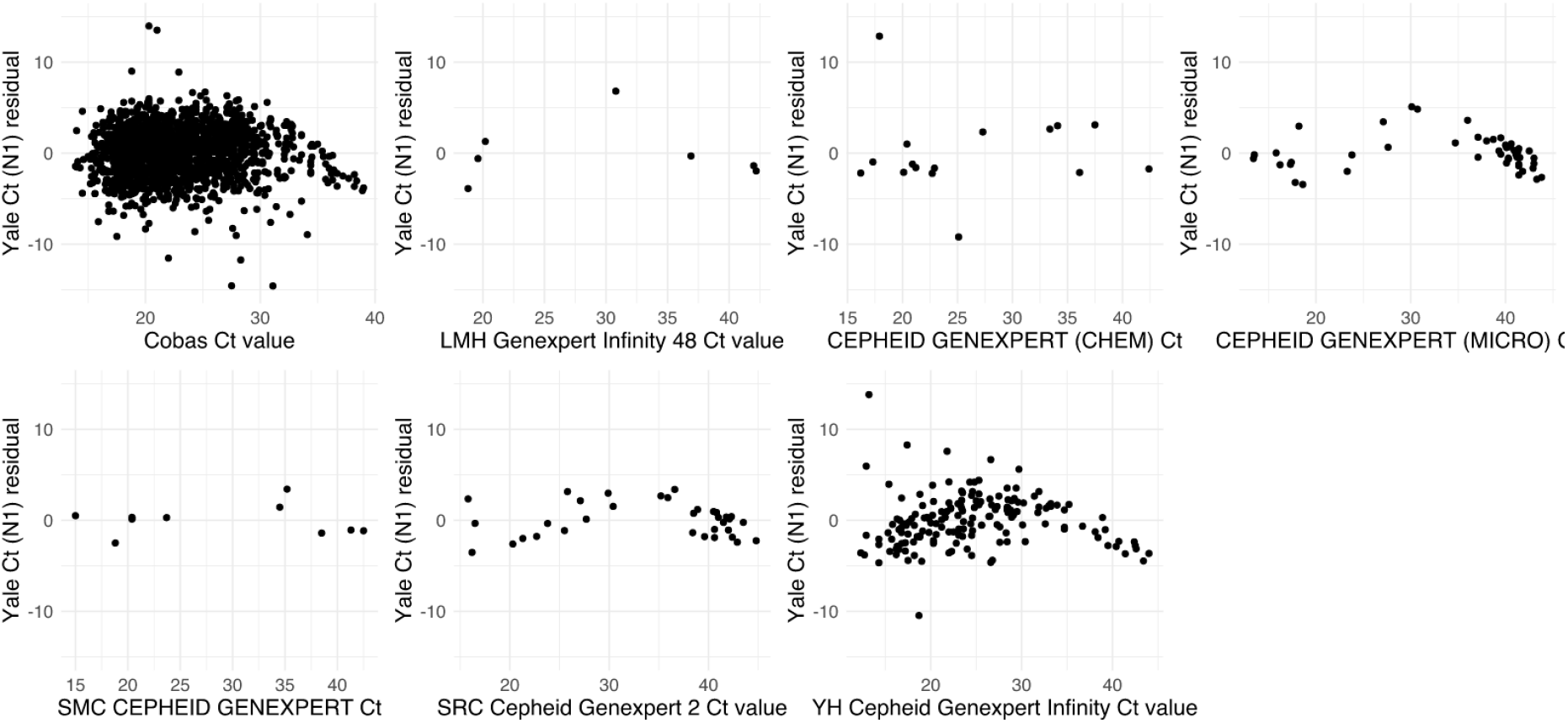
Residuals from Ct regressions for each platform.

## Notes

### Competing Interest Statement

The authors have declared no competing interest.

### Funding Statement

This study was funded by research grants from Dr. Akiko Iwasaki.

### Author Declarations

Ethics committee/IRB of Yale University gave ethical approval for this work.

### Summary of Updates

- Correction of text. - Correction of y-axis label in Figure 1c.

